# Are population-wide mass media campaigns still relevant in an era of low smoking prevalence? Insights from the *Power to Quit* campaign in England

**DOI:** 10.64898/2026.09.14.26362985

**Authors:** Sarah E. Jackson, Jamie Brown

## Abstract

**Introduction:** The role of population-wide smoking cessation campaigns is uncertain as smoking prevalence reaches historically low levels. We examined whether the *Power to Quit* campaign in England, which aimed to increase uptake of evidence-based cessation support, particularly digital aids, was associated with changes in quitting behaviour.

**Methods:** We analysed nationally-representative cross-sectional surveys of adults in England who smoked in the past year, conducted immediately before (*n*=803) and after (*n*=1,713) the campaign (February-April 2026). Logistic regression estimated associations with quit attempts, digital cessation support, local Stop Smoking Services (SSS), and quit success, overall and by social grade. Exploratory analyses examined participants recognising campaign materials (*n*=799). Smoking Toolkit Study (STS) data (January 2023-June 2026; *n*=11,442) were used to contextualise findings against longer-term trends.

**Results:** In the primary pre/post analysis, there were increases in quit attempts (48.4% to 52.3%, aOR=1.16 [95%CI=0.97-1.38]), attempts involving digital support (16.0% to 18.4%, aOR=1.18 [0.92-1.50], and attempts involving SSS (2.4% to 3.6%, aOR=1.53 [0.89-2.65]), although estimates were uncertain. Associations were larger among participants recognising campaign materials (quit attempts: 64.9%, aOR=1.82 [1.47-2.26]; digital support: 28.7%, aOR=1.88 [1.44-2.45]; SSS: 5.4%, aOR=2.14 [1.19- 3.88]). Increases in support uptake were particularly evident among less advantaged adults. There was no clear evidence of a change in quit success. STS data showed increases in support uptake continued from before the campaign.

**Conclusions:** *Power to Quit* coincided with increased engagement with cessation support, particularly among less advantaged adults and those recognising campaign materials. Increases in support continued pre-campaign trends, indicating that the contribution of the campaign occurred within a broader period of changing cessation behaviour.

**Implications:** National mass media campaigns appear to still contribute to increasing engagement with evidence-based cessation support when smoking prevalence is low, particularly among more disadvantaged groups, where smoking remains more common and quit success rates are lower. Although *Power to Quit* was a population-wide campaign, media planning prioritised areas with higher smoking prevalence and settings where smoking was more common and habitual, alongside sociodemographic targeting where feasible. These findings suggest that population-wide campaigns and targeted delivery strategies can be complementary rather than competing approaches.

## Introduction

Mass media tobacco control campaigns are an effective population-level intervention for increasing smoking cessation activity.^1^ However, in a context where smoking prevalence in England has reached historically low levels,^2,3^ it is increasingly important to consider the role of broad population campaigns within tobacco control strategies. Key questions include whether population-wide campaigns remain an efficient use of public health resources, or whether investment should instead prioritise local interventions for groups experiencing the greatest burden of smoking-related harm. This question is particularly relevant because smoking is increasingly concentrated among people experiencing socioeconomic disadvantage and remains a major contributor to health inequalities in England.^2,4,5^

In England, most people who smoke report wanting to quit^6^ and many have made previous quit attempts.^7^ However, most quit attempts continue to occur without evidence-based support.^8^ This represents an important opportunity for improving cessation outcomes, as quit attempts supported by behavioural or pharmacological interventions are substantially more likely to succeed than unaided attempts.^8–10^

The UK Department of Health and Social Care (DHSC) ran the *Power to Quit* campaign^11^ between 16 February and 5 April 2026. This national smoking cessation mass media campaign aimed to improve the quality of quit attempts by encouraging people who smoke to use effective cessation support, particularly digital resources such as the NHS Quit Smoking app and Personal Quit Plan. The campaign positioned these tools as an enhancement to, rather than a substitute for, people’s own motivation and determination to quit. Although *Power to Quit* was a population-wide campaign, media planning prioritised geographic areas with higher smoking prevalence and environments in which smoking is more prevalent or habitual, alongside sociodemographic targeting where this was possible across different advertising channels. The campaign was launched during a period of increased government investment in local Stop Smoking Services,^12^ with additional funding introduced from April 2024 to expand access to evidence-based behavioural and pharmacological support for people who smoke.

This study aimed to examine the role that population-wide mass media campaigns can play in supporting smoking cessation in an era of low smoking prevalence and alongside expanded provision of targeted cessation support. Specifically, we evaluated whether the *Power to Quit* campaign was associated with quit attempts, use of cessation support during quit attempts, including digital cessation tools and local Stop Smoking Services, and quit success. Given the campaign’s emphasis on reaching communities with higher smoking prevalence, we also explored whether these associations differed between adults from more advantaged (ABC1) and less advantaged (C2DE) occupational social grades.

## Methods

### Data sources

We used two data sources. First, two cross-sectional surveys commissioned by DHSC and conducted by market research company Ipsos immediately before (4–11 February 2026) and after (9–23 April 2026) the *Power to Quit* campaign. A new sample was recruited at each wave, enabling comparison of population-level measures before and after the campaign. Samples were recruited using quotas for age, gender, region, and social grade to reflect the adult population in England, and survey weights were applied to align with population distributions. The same blend of online access panels was used at each wave, with similar contributions from each panel to support comparability over time.

Second, data from the Smoking Toolkit Study,^13,14^ were included to contextualise observed changes against longer-term trends. This is an ongoing monthly cross-sectional household survey commissioned by UCL and funded by Cancer Research UK, which is also conducted by Ipsos. It uses a hybrid of random probability and quota sampling to recruit a new sample of approximately 1,700 adults in England each month, with data collected through telephone interviews. Survey weights are generated using raking (iterative proportional fitting) to align the sample with the population distribution of England on key sociodemographic characteristics and smoking-related variables. We included Smoking Toolkit Study data from January 2023 to June 2026.

### Participants

We analysed data from adults aged ≥18 years living in England who reported smoking cigarettes or other tobacco products within the past 12 months and reported whether they had made a quit attempt over this period. This provided total samples of 2,516 in the DHSC-commissioned surveys (803 pre-campaign and 1,713 post-campaign) and 11,442 in the Smoking Toolkit Study.

### Measures

We analysed data on quit attempts, use of digital cessation support, use of local Stop Smoking Services, and quit success. Outcomes were derived separately for DHSC-commissioned pre- and post-campaign surveys and the Smoking Toolkit Study, using harmonised definitions where possible (see **Supplementary File 1**).

In each survey, participants were classified as having made a past-year quit attempt if they reported at least one attempt to stop smoking during the previous 12 months. Those who reported a past-year quit attempt were then asked follow-up questions about the types of support used in the most recent attempt and whether the attempt was successful. Digital support included use of NHS digital cessation resources (including the NHS Better Health Quit Smoking website, NHS Quit Smoking app, and NHS Personal Quit Plan), other websites, and other stop smoking apps. Use of local Stop Smoking Services included face-to-face or online sessions. Successful cessation was defined as the participant reporting that they were still not smoking following the most recent quit attempt.

Sociodemographic characteristics included age (18-24, 25-34, 35-44, 45-54, 55-64 and S65 years), gender (man, woman, or another gender), and occupational social grade (ABC1 = managerial, professional, and upper supervisory occupations vs. C2DE = manual routine, semi-routine, lower supervisory, state pension, and long-term unemployed), classified according to the National Readership Survey social grade classification.^15^ Recognition of *Power to Quit* campaign materials was also assessed in the DHSC-commissioned post-campaign survey.

### Statistical analysis

Data were analysed on complete cases using R v.4.6.0. For each data source, outcomes were analysed using two denominators: (1) all adults who smoked in the past year, and (2) the subset of adults who reported a past-year quit attempt.

DHSC-commissioned survey data were summarised as proportions before and after the campaign. Logistic regression models estimated associations between survey wave (pre-campaign vs post-campaign) and each outcome, adjusted for age, gender, and occupational social grade. Additional analyses were stratified by occupational social grade (ABC1 vs C2DE) to explore whether associations differed between more and less advantaged groups. Primary analyses included all eligible participants. In exploratory analyses, we compared the pre-campaign sample with the subset of post-campaign participants who reported recognising the campaign materials, to examine whether associations were stronger among those exposed to the campaign. Regression results are reported as adjusted odds ratios (aORs) with 95% confidence intervals (CIs).

Smoking Toolkit Study data were analysed descriptively using weighted three-month right-aligned moving averages and 95% CIs for each outcome, calculated using data from January 2023 to June 2026 and plotted from March 2023 onwards, to contextualise the pre/post survey findings against longer-term trends. To provide a simpler summary of changes over time and reduce the influence of month-to-month variability, we also calculated the weighted prevalence (with 95% CIs) of each outcome within January–June and July–December of each year over the same period. Additional descriptive analyses stratified these trends by occupational social grade (ABC1 vs C2DE) for adults who smoked in the past year.

In line with contemporary recommendations for statistical inference, we focused on estimation of effect sizes and their uncertainty rather than dichotomous interpretation of statistical significance based on *p* values.^16,17^ Associations are therefore described in terms of effect sizes, directions of effect, and uncertainty reflected by 95% CIs.

## Results

### Participant characteristics

The pre-campaign survey included 803 adults who had smoked in the past year, of whom 383 reported making a past-year quit attempt. The post-campaign survey included 1,713 adults who had smoked in the past year, of whom 901 had made a past-year quit attempt. Among post-campaign respondents, 799 adults who had smoked in the past year and 518 who had attempted to quit reported recognising the *Power to Quit* campaign materials and were included in exploratory analyses. The demographic composition of the two survey waves was similar, with no substantial differences in age, gender, or occupational social grade (**Table S1**).

The Smoking Toolkit Study sample included 11,442 adults who had smoked in the past year, of whom 4,091 reported a past-year quit attempt; characteristics of this sample are provided in **Table S2**.

### Pre- and post-campaign surveys

**Table 1** summarises quitting outcomes before and after the *Power to Quit* campaign. In the primary pre/post analysis, there was some evidence of increases in quit attempts and use of cessation support following the campaign, although estimates were generally imprecise. Among adults who smoked in the past year, there was a small, uncertain increase in the proportion reporting a quit attempt, from 48.4% before the campaign to 52.3% after the campaign (aOR 1.16, 95% CI 0.97– 1.38). The proportion reporting a quit attempt involving digital support increased modestly from 16.0% to 18.4% (aOR 1.18, 95% CI 0.92–1.50), while the point estimate for quit attempts involving local Stop Smoking Services suggested a larger increase but was less precise (2.4% to 3.6%; aOR 1.53, 95% CI 0.89–2.65). Among adults who attempted to quit, increases in support use were smaller, with digital support increasing from 33.1% to 35.1% and local Stop Smoking Services from 4.9% to 6.8%. There was no evidence of a change in quit success following the campaign, either among all adults who smoked in the past year or among those who attempted to quit.

**Table 1.**
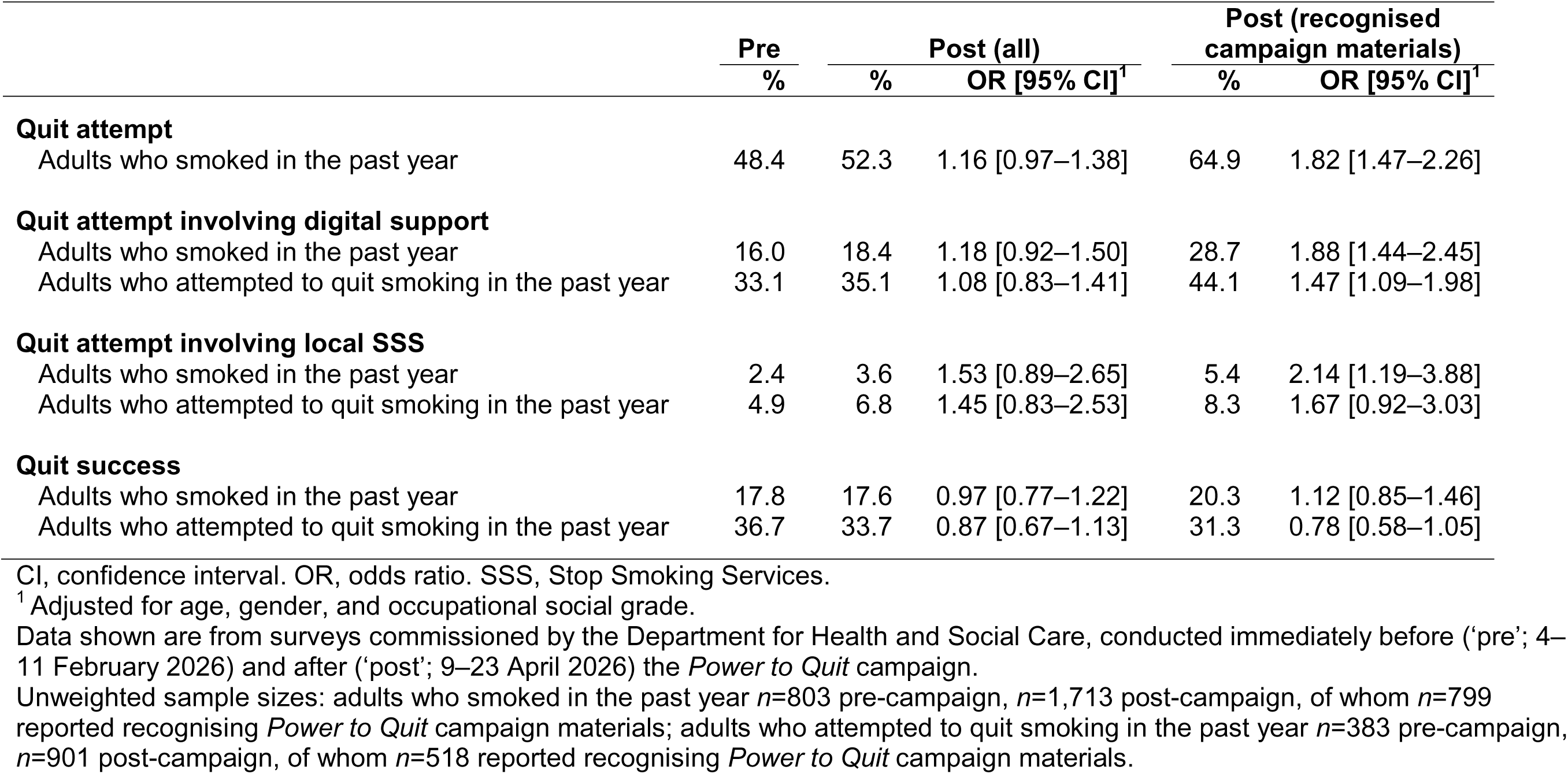
Quitting outcomes before and after the *Power to Quit* campaign, overall and among participants recognising campaign materials.

In exploratory analyses restricting the post-campaign sample to participants who reported recognising the *Power to Quit* campaign materials, associations were larger than in the primary pre/post analysis (**Table 1**). Among adults who smoked in the past year, the proportion reporting a quit attempt increased from 48.4% before the campaign to 64.9% among those recognising campaign materials after the campaign (aOR 1.82, 95% CI 1.47–2.26). The proportion reporting quit attempts involving digital support increased from 16.0% to 28.7% (aOR 1.88, 95% CI 1.44– 2.45) and quit attempts involving local Stop Smoking Services increased from 2.4% to 5.4% (aOR 2.14, 95% CI 1.19–3.88). Among adults who attempted to quit, use of digital support increased from 33.1% to 44.1% (aOR 1.47, 95% CI 1.09–1.98), while use of local Stop Smoking Services increased from 4.9% to 8.3%, although this estimate was less precise (aOR 1.67, 95% CI 0.92– 3.03). There was no clear evidence of an association between campaign recognition and quit success.

Stratified analyses by occupational social grade are presented in **Table 2**. In the primary analysis, increases in quit attempts and cessation support uptake were concentrated among less advantaged (C2DE) adults. Among C2DE adults who smoked in the past year, quit attempts increased from 44.7% to 51.5% (aOR 1.31, 95% CI 1.02–1.70), whereas estimates among more advantaged (ABC1) adults remained similar (51.5% to 53.0%; aOR 1.06, 95% CI 0.83–1.35). Quit attempts involving digital support showed a similar pattern, increasing among C2DE adults who smoked in the past year (9.9% to 16.1%; aOR 1.73, 95% CI 1.16–2.57) but remaining similar among ABC1 adults (21.2% to 20.1%; aOR 0.94, 95% CI 0.69–1.28), with a consistent pattern observed among those who attempted to quit (C2DE: 22.2% to 31.4%; aOR 1.54, 95% CI 1.00– 2.36). Estimates for local Stop Smoking Services were directionally similar but less precise. There was little evidence of differences in quit success by occupational social grade.

**Table 2.**
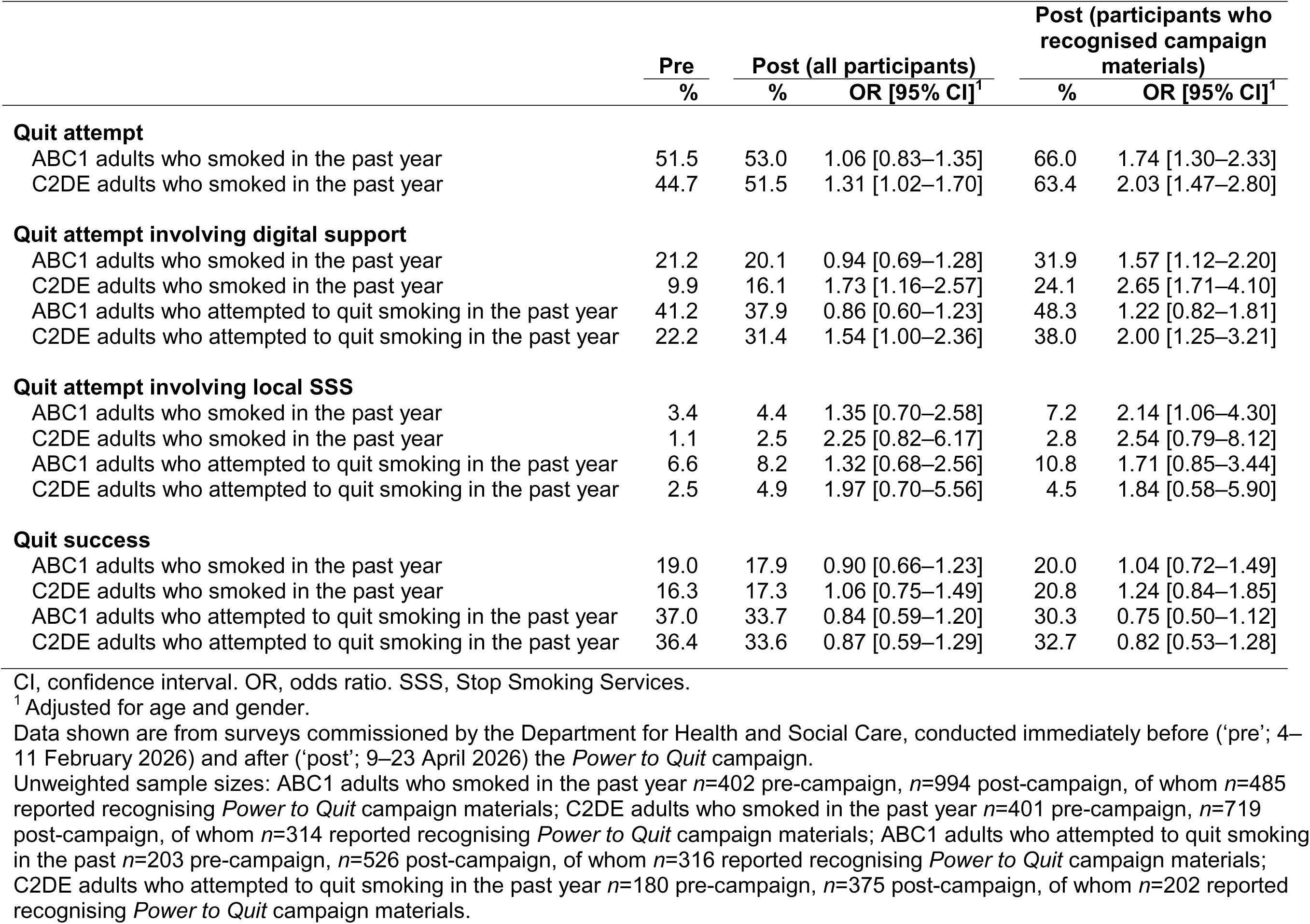
Quitting outcomes before and after the *Power to Quit* campaign by occupational social grade, overall and among participants recognising campaign materials.

Patterns by social grade were broadly similar in analyses restricted to participants who recognised campaign materials, although increases in digital support appeared particularly pronounced among less advantaged adults (**Table 2**). Among C2DE adults who smoked in the past year, the proportion reporting quit attempts involving digital support increased from 9.9% before the campaign to 24.1% among those recognising campaign materials (aOR 2.65, 95% CI 1.71–4.10).

### Longer-term trends (Smoking Toolkit Study)

**Figures 1 and 2** show longer-term trends in quitting outcomes in England, with corresponding six- month prevalence estimates presented in **Tables S3–S6**.

**Figure 1.**
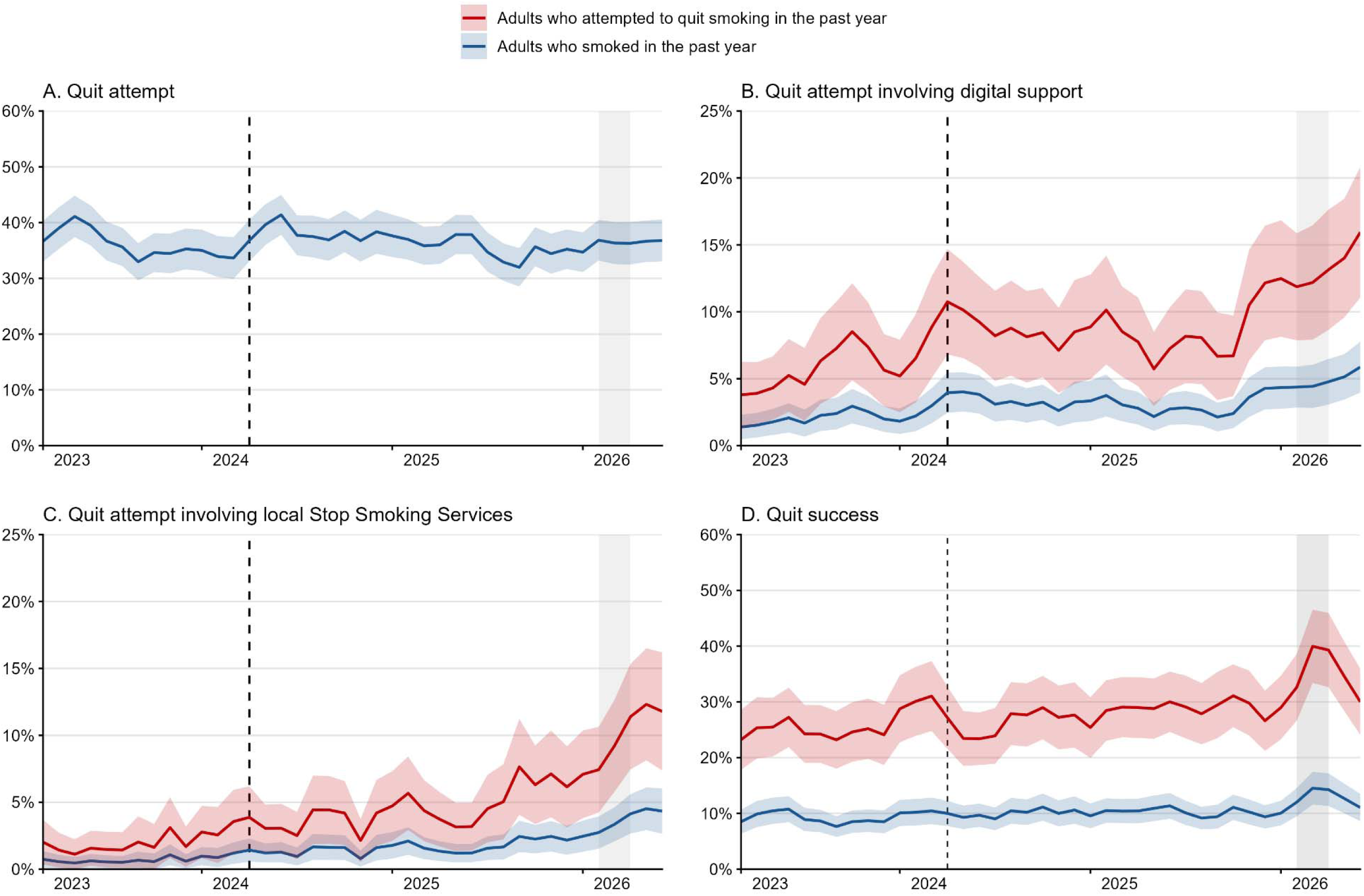
Longer-term trends in quit attempts, use of support, and quit success in England, 2023–2026. Data shown are weighted three-month moving averages from the Smoking Toolkit Study, calculated using data from January 2023 to June 2026 and plotted from March 2023 onwards. Lines indicate means and shaded bands 95% confidence intervals. Dashed vertical lines indicate the expansion of government investment in local Stop Smoking Services (April 2024). Shaded areas indicate the period of the *Power to Quit* campaign (February–April 2026). Unweighted sample sizes: adults who smoked in the past year *n*=11,442; adults who attempted to quit smoking in the past year *n*=4,091.

**Figure 2.**
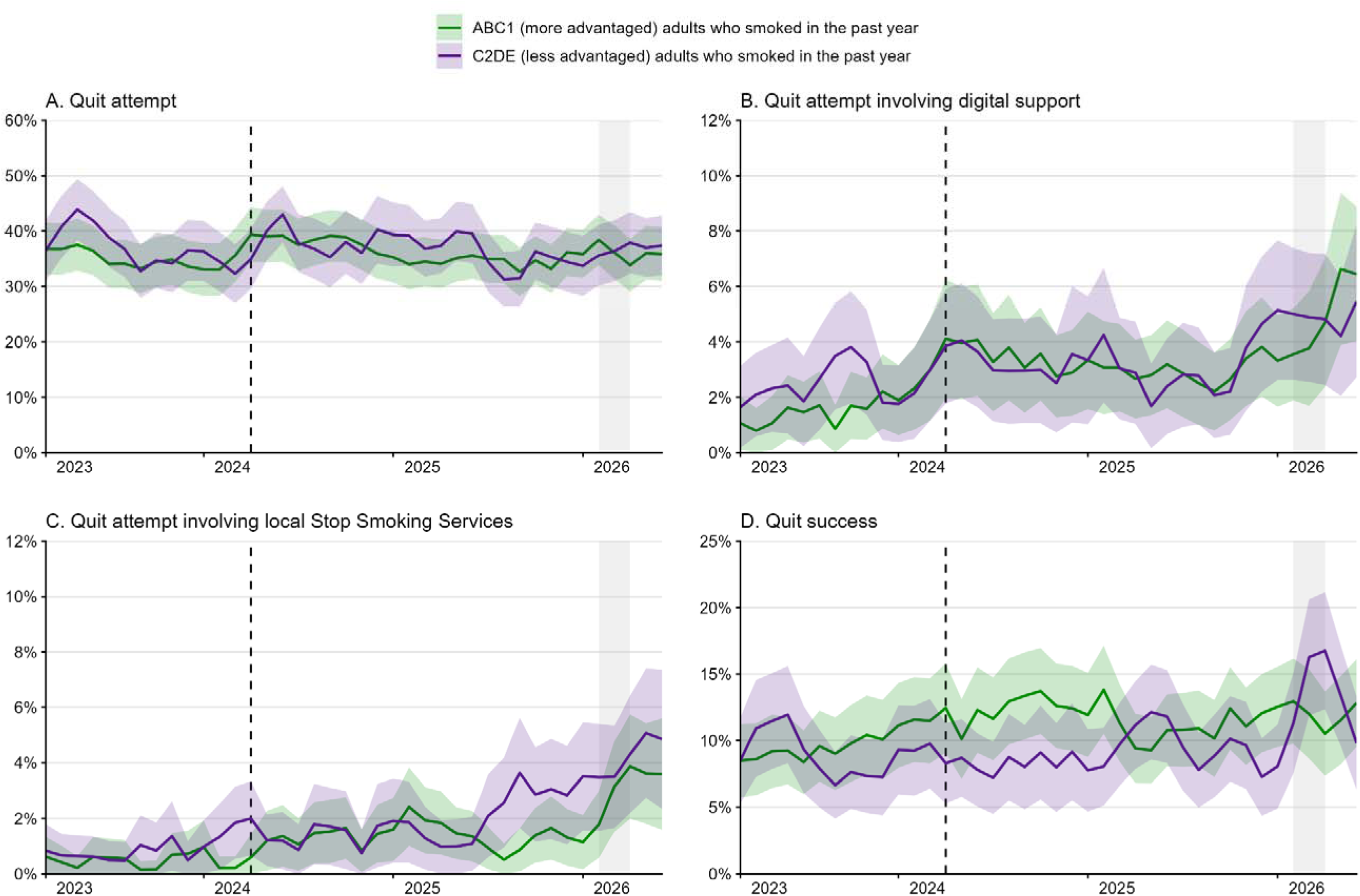
Longer-term trends in quit attempts, use of support, and quit success in England, 2023–2026 – by occupational social grade. Data shown are weighted three-month moving averages from the Smoking Toolkit Study, calculated using data from January 2023 to June 2026 and plotted from March 2023 onwards. Lines indicate means and shaded bands 95% confidence intervals. Dashed vertical lines indicate the expansion of government investment in local Stop Smoking Services (April 2024). Shaded areas indicate the period of the *Power to Quit* campaign (February–April 2026). Unweighted sample sizes: ABC1 adults who smoked in the past year *n*=6,510; C2DE adults who smoked in the past year *n*=4,932.

The prevalence of quit attempts among adults who smoked in the past year was relatively stable over time, with three-month moving averages fluctuating between 32.0% [95%CI 28.5–35.4] and 41.4% [37.8–45.0], and no clear change coinciding with the *Power to Quit* campaign period (**Figure 1A**). In stratified analyses by occupational social grade, estimates were in the direction of a small increase in quit attempts among less advantaged (C2DE) adults, from 33.7% [28.3–39.2] immediately before the campaign in January 2026 to 37.4% [32.0–42.8] in June 2026, while estimates among more advantaged (ABC1) adults remained similar (35.9% [31.6–40.2] to 35.9% [31.1–40.7]; **Figure 2A**), broadly consistent with the pre- and post-campaign analysis. However, similar fluctuations were observed earlier in the time series, meaning these patterns should be interpreted cautiously and may reflect underlying variability rather than a campaign-related change.

Uptake of cessation support increased from mid to late 2025 onwards, continuing an upward trend that began before the campaign period. Among adults who smoked in the past year, the proportion reporting a quit attempt involving digital support increased from 1.4% [0.5–2.3] in March 2023 to 4.3% [2.8–5.9] immediately before the campaign in January 2026, and further increased to 5.9% [4.0–7.8] by June 2026 (**Figure 1B**). Consistent with this pattern, six-month estimates increased from 3.2% [2.3–4.2] in July–December 2025 to 5.1% [3.9–6.4] in January–June 2026 (**Table S4**). Among adults who made a quit attempt, use of digital support increased from 3.8% [1.3–6.3] in March 2023 to 12.5% [8.1–16.8] before the campaign, and to 15.9% [11.1–20.8] by June 2026 (**Figure 1B**). Six-month estimates similarly increased from 9.6% [6.9–12.4] in July–December 2025 to 14.1% [10.8–17.3] in January–June 2026 (**Table S4**). Increases were observed across occupational social grades (**Table S4**), although the rise among C2DE adults appeared to begin before the campaign period, suggesting that these patterns may reflect broader changes in support uptake rather than a campaign-specific effect (**Figure 2B**).

The proportion reporting a quit attempt involving local Stop Smoking Services showed a similar pattern, increasing from 0.7% [0.1–1.4] of adults who smoked in the past year in March 2023 to 2.5% [1.3–3.6] immediately before the campaign and 4.3% [2.7–6.0] by June 2026 (**Figure 1C**). Six-month estimates increased from 2.3% [1.5–3.1] in July–December 2025 to 3.8% [2.8–4.9] in January–June 2026 (**Table S5**). Among adults who attempted to quit, use of local Stop Smoking Services increased from 2.0% [0.4–3.7] to 7.1% [3.8–10.4] before the campaign, and to 11.8% [7.4–16.2] by June 2026 (**Figure 1C**). In stratified analyses, increases were observed among both ABC1 and C2DE adults (**Table S5**), although the timing differed: among C2DE adults, uptake increased earlier, with a notable rise during the latter half of 2025, whereas the increase among ABC1 adults was concentrated later, during the campaign period (**Figure 2C**).

Quit success showed short-term increases around the campaign period. Among adults who smoked in the past year, the proportion reporting a successful quit attempt rose from an average of 10.2% before the campaign (10.1% [7.9–12.3] in January 2026) to a peak of 14.5% [11.6–17.4] in March 2026, before declining to 11.0% [8.6–13.5] by June 2026 (**Figure 1D**). Six-month estimates increased from 9.4% [7.9–10.9] in July–December 2025 to 12.8% [10.9–14.7] in January–June 2026 (**Table S6**). Among adults who made a quit attempt, quit success increased from an average of 28.1% before the campaign (29.8% [24.0–35.6] in January 2026) to a peak of 40.0% [33.4–46.5], before declining to 30.0% [24.1–36.0] by June 2026 (**Figure 1D**). In stratified analyses, the increase appeared more pronounced among less advantaged (C2DE) adults, among whom the proportion reporting a successful quit attempt increased from an average of 9.5% before the campaign (8.1% [4.9–11.2] in January 2026) to a peak of 16.8% [12.4–21.2] in April 2026, before declining to 9.8% [6.3–13.3] by June 2026 (**Figure 2D**). In contrast, quit success among more advantaged (ABC1) adults remained relatively stable over the same period (average pre-campaign = 11.1%; range January – June 2025 = 10.5–12.9%; **Figure 2D**).

## Discussion

In this evaluation of the national *Power to Quit* smoking cessation mass media campaign in England, we examined changes in quitting behaviours and use of cessation support using complementary data sources: contemporaneous pre- and post-campaign surveys and longer-term data from the Smoking Toolkit Study.

In the primary pre/post analysis, there was some evidence of increases in quit attempts and use of cessation support following the campaign, although estimates were generally uncertain among all adult smokers. Increases in quit attempts and support uptake were evident among less advantaged adults. Exploratory analyses restricted to post-campaign participants who recognised campaign materials showed larger increases across several outcomes, including quit attempts, digital cessation support, and use of local Stop Smoking Services. These larger associations are consistent with a potential contribution of the campaign to changes in cessation behaviour.

However, this analysis should be interpreted cautiously because people who recognised campaign materials may have differed from those who did not in ways that also influenced their likelihood of attempting to quit or using cessation support.

The pattern of changes was broadly aligned with the intended mechanism of *Power to Quit*: encouraging people who smoke to engage with evidence-based support as part of a quit attempt. Increases were observed across different forms of support, including digital resources and local Stop Smoking Services, suggesting that promoting digital cessation tools did not displace other forms of support. Digital tools may be particularly valuable within population-level cessation strategies because they can provide accessible and scalable support to large numbers of people who smoke, while also complementing more intensive behavioural support delivered through local services.^18^ The campaign may therefore have contributed to a broader shift towards supported quitting rather than simply increasing awareness of a single intervention. This pattern is consistent with previous evidence that mass media campaigns are more likely to influence proximal cessation behaviours, such as motivation to quit, quit attempts, and engagement with cessation support, than longer-term cessation outcomes, which are influenced by multiple subsequent factors.^1,19,20^ Increasing the proportion of quit attempts that involve evidence-based support remains an important route to improving cessation outcomes.^8–10^

The Smoking Toolkit Study findings provide important context for interpreting the pre/post survey results. Uptake of digital cessation support and local Stop Smoking Services increased during late 2025 and early 2026, but these increases were already underway before the *Power to Quit* campaign period. This suggests that the campaign took place within a broader period of increasing engagement with cessation support rather than being the sole driver of these changes. The timing of the *Power to Quit* campaign coincided with wider tobacco control activity, including increased investment in local Stop Smoking Services^12^ and other public health initiatives. This included a separate national campaign from December 2025 to April 2026 promoting the NHS Healthy Choices Quiz,^21^ which provides personalised advice on health behaviours, including signposting people who smoke towards cessation support. These activities may have contributed to changes in cessation behaviours, and potential synergies between mass media campaigns, public health messaging, and expanded service provision may have strengthened their collective impact. Such overlap reflects the way population-level tobacco control operates in practice, where multiple interventions are delivered together rather than in isolation. Evaluating mass media campaigns as standalone interventions may therefore underestimate their contribution, as their intended function is often to increase demand for and engagement with services that enable behaviour change.

There was less evidence that the campaign was associated with changes in quit success. In the pre/post survey analysis, quit success was similar before and after the campaign, both among all adults who smoked in the past year and among those who attempted to quit. The Smoking Toolkit Study showed some short-term increases in quit success around the campaign period, but similar fluctuations were observed earlier in the time series. Longer-term follow-up will therefore be important to determine whether increased engagement with cessation support translates into sustained improvements in cessation outcomes.

The findings also provide insights into the potential role of population-wide campaigns in addressing inequalities in smoking. Although *Power to Quit* was delivered as a population-wide campaign, its media strategy prioritised areas with higher smoking prevalence and settings where smoking was more common or habitual, alongside sociodemographic targeting where feasible. Consistent with this targeting, increases in quit attempts and support uptake appeared particularly evident among less advantaged adults. While these subgroup analyses require cautious interpretation, they suggest that broad-reach campaigns can potentially be designed and delivered in ways that reach groups experiencing the greatest burden of smoking. Previous evidence indicates that the extent to which mass media campaigns reduce inequalities may depend less on whether campaigns are universally delivered or specifically targeted, and more on achieving sufficient reach and exposure among disadvantaged groups.^1^ Targeted approaches can increase message relevance, but may also reduce overall campaign exposure if resources are spread across multiple tailored messages.^1^ Our findings therefore highlight the potential for combining population-level approaches with targeted delivery strategies, rather than viewing these as competing alternatives.

This study has several strengths. It used nationally representative samples and triangulated findings across two complementary data sources: evaluation-specific pre/post campaign surveys and an independent monthly surveillance system capturing longer-term trends in smoking and cessation behaviours. The inclusion of multiple outcomes allowed assessment of changes across different stages of the quitting process, from attempting to quit through engagement with support and subsequent cessation.

There were also limitations. First, the observational pre/post design means that changes in cessation behaviours cannot be causally attributed to the *Power to Quit* campaign. The campaign occurred alongside other tobacco control activities and wider changes in cessation support provision, meaning that the independent contribution of the campaign cannot be isolated. Second, although the Smoking Toolkit Study provided valuable contextual information, the use of three-month moving averages may have reduced sensitivity to short-term changes occurring around the campaign period. Third, all measures were based on self-report and may be subject to recall or reporting biases. Fourth, outcomes were based on past-year quitting activity, meaning that some outcomes may reflect behaviours that occurred before the survey date and therefore may not align precisely with the timing of campaign exposure. However, this lag would be expected to apply similarly across survey waves and therefore should not substantially affect the assessment of changes in trends over time. Fifth, there were some differences in outcome assessment between the DHSC-commissioned surveys and Smoking Toolkit Study, which may contribute to differences in patterns of results. For example, the DHSC-commissioned surveys captured specific NHS digital cessation resources, including the NHS Quit Smoking app and Personal Quit Plan, whereas the Smoking Toolkit Study measure did not distinguish these resources separately and may therefore capture a narrower range of digital support use. Finally, some estimates were imprecise because smoking and use of some forms of support are now relatively uncommon in the population.

Overall, these findings suggest that mass media campaigns can continue to play a role within comprehensive tobacco control strategies by encouraging engagement with cessation support. In combination with accessible cessation services and wider tobacco control measures, campaigns may help support further reductions in smoking prevalence and inequalities.

## Data Availability

All data produced in the present study are available upon reasonable request to the authors

## Declarations

### Data availability

Data are available from the corresponding author on reasonable request.

### Ethics approval

Ethical approval for the Smoking Toolkit Study was granted originally by the UCL Ethics Committee (ID 0498/001). Participants provide informed consent to take part in the study, and all methods are carried out in accordance with relevant regulations. The data are not collected by UCL and are anonymised when received by UCL. Ethical approval for secondary analysis of the DHSC- commissioned pre- and post-campaign surveys was also granted by the UCL Ethics Committee (ID 5034).

## References

1. Durkin, S., Brennan, E. & Wakefield, M. Mass media campaigns to promote smoking cessation among adults: an integrative review. Tob. Control 21, 127–138 (2012).

2. Office for National Statistics. Adult smoking habits in the UK: 2024. https://www.ons.gov.uk/peoplepopulationandcommunity/healthandsocialcare/healthandlifeexpectancies/bulletins/adultsmokinghabitsingreatbritain/2024 (2025).

3. Jackson, S. E., Shahab, L. & Brown, J. The end of smoking in England? The importance of considering different metrics of success. Tob. Control https://doi.org/10.1136/tc-2025-059469 (2025) doi:10.1136/tc-2025-059469.

4. Theodoulou, A., Hartmann-Boyce, J., Lindson, N., Fanshawe, T. R. & Jackson, S. E. Smoking and Quitting Behaviors by Different Indicators of Socioeconomic Position in England: A Population Study, 2014 to 2023. Nicotine Tob. Res. ntag003 (2026) doi:10.1093/ntr/ntag003.

5. Action on Smoking and Health. Health Inequalities Resource Pack. Action on Smoking and Health https://ash.org.uk/ash-local-toolkit/health-inequalities-resource-pack/ (2019).

6. Jackson, S. E., Brown, J., Shahab, L. & Cox, S. Associations between non-daily smoking and motivation to stop smoking: A population study in England 2021–2024. Addiction 120, 2519– 2526 (2025).

7. Jackson, S. E., Tattan-Birch, H., Shahab, L., Beard, E. & Brown, J. Have there been sustained impacts of the COVID-19 pandemic on trends in smoking prevalence, uptake, quitting, use of treatment, and relapse? A monthly population study in England, 2017–2022. BMC Med. 21, 474 (2023).

8. Jackson, S. E., Brown, J., Buss, V. & Shahab, L. Prevalence of Popular Smoking Cessation Aids in England and Associations With Quit Success. *JAMA Netw*. Open 8, e2454962 (2025).

9. Hartmann-Boyce, J. et al. Behavioural interventions for smoking cessation: an overview and network meta-analysis. Cochrane Database Syst. Rev. https://doi.org/10.1002/14651858.CD013229.pub2 (2021) doi:10.1002/14651858.CD013229.pub2.

10. Lindson, N. et al. Pharmacological and electronic cigarette interventions for smoking cessation in adults: component network meta-analyses. Cochrane Database Syst. Rev. 10.1002/14651858.CD015226.pub2 (2023) doi:10.1002/14651858.CD015226.pub2.

11. Department for Health and Social Care. The Power to Quit Smoking. Campaign Resource Centre https://campaignresources.dhsc.gov.uk/campaigns/quit-smoking-national-campaign-2026/ (2026).

12. Department of Health and Social Care. Stopping the Start: Our New Plan to Create a Smokefree Generation. https://www.gov.uk/government/publications/stopping-the-start-our-new-plan-to-create-a-smokefree-generation (2023).

13. Fidler, J. A. et al. ‘The smoking toolkit study’: a national study of smoking and smoking cessation in England. BMC Public Health 11, 479 (2011).

14. Kock, L. et al. Protocol for expansion of an existing national monthly survey of smoking behaviour and alcohol use in England to Scotland and Wales: The Smoking and Alcohol Toolkit Study. Wellcome Open Res. 6, 67 (2021).

15. National Readership Survey. Social grade - definitions and discriminatory power. (2007).

16. Cumming, G. The New Statistics: Why and How. Psychol. Sci. 25, 7–29 (2014).

17. Calin-Jageman, R. J. & Cumming, G. The New Statistics for Better Science: Ask How Much, How Uncertain, and What Else Is Known. Am. Stat. 73, 271–280 (2019).

18. Li, S. et al. Efficacy of digital interventions for smoking cessation by type and method: a systematic review and network meta-analysis. *Nat*. Hum. Behav. 9, 2054–2065 (2025).

19. Wakefield, M. A., Loken, B. & Hornik, R. C. Use of mass media campaigns to change health behaviour. Lancet 376, 1261–1271 (2010).

20. Bala, M. M., Strzeszynski, L. & Topor-Madry, R. Mass media interventions for smoking cessation in adults. Cochrane Database Syst. Rev. https://doi.org/10.1002/14651858.CD004704.pub4 (2017) doi:10.1002/14651858.CD004704.pub4.

21. NHS. Healthy Choices Quiz. (2026).

