## Supplementary file 1 for "Are population-wide mass media campaigns still relevant in an era of low smoking prevalence? Insights from the *Power to Quit* campaign in England"

**Measures**

**Quit attempts**

| Base: all adults who smoked in the past year | | |
|---|---|---|
| Survey | **Ipsos pre/post campaign** | **Smoking Toolkit Study** |
| Variable name | F1 | Q632b7 |
| Item wording and response options | Have you made an attempt to quit smoking in the past 12 months?<br>By quit attempt, we mean stopping smoking cigarettes or other tobacco products (e.g. pipe, cigar, shisha, etc.), though you may still be using a vape (electronic cigarette or vaping device).<br><br>24. Yes - and I am still not smoking cigarettes<br>25. Yes - but I have started smoking cigarettes again<br>26. No - I have not made a quit attempt<br>27. Don't know | How many serious attempts to stop smoking have you made in the last 12 months? By serious attempt I mean you decided that you would try to make sure you never smoked again. Please include any attempt that you are currently making and please include any successful attempt made within the last year.<br><br>ALLOW NUMERIC RANGE 0-150, DK |
| Coding: past-year quit attempt | 1-2 = yes<br>3 = no<br>4 = NA | 1+ = yes<br>0 = no<br>Don't know = NA |

**Use of support in the most recent quit attempt**

| Base: adults who smoked in the past year and made at least one past-year quit attempt | | |
|---|---|---|
| Survey | **Ipsos pre/post campaign** | **Smoking Toolkit Study** |
| Variable name | F4 | Q632e40 |
| Item wording and response options | Which, if any, of the following did you try to help you stop smoking cigarettes or other tobacco products during your most recent quit attempt?<br><br>1. Used Nicotine replacement products (for example, patches, gum, inhaler)<br>2. Took Zyban (bupropion), Champix (varenicline) or other pills/tablets on prescription<br>3. Used tobacco-free nicotine pouch/pod or 'white pouches' (e.g., Zyn, On!, Nordic Spirit, Velo, Lyft, Skruf)<br>4. Used Juul<br>5. Used heat-not-burn cigarettes (e.g. iQOS with HEETS, heatsticks)<br>6. Used disposable e-cigarettes/vapes<br>7. Used reusable e-cigarettes/vapes<br>8. Used the NHS Better Health Quit Smoking website<br>9. Used another online website<br>10. Used the NHS Quit Smoking app<br>11. Used another stop smoking app<br>12. Used the NHS Personal Quit Plan tool/quiz<br>13. Joined the NHS quit smoking email support programme<br>14. Called the National Smokefree Helpline<br>15. Spoke to my GP or other healthcare professional | Which, if any, of the following did you try to help you stop smoking during the most recent serious quit attempt?<br><br>1. Nicotine replacement product (eg. patches\gum\inhaler) without a prescription<br>2. Nicotine replacement product on prescription or given to you by a health professional<br>3. Zyban (bupropion)<br>4. Champix (varenicline)<br>5. Attended a Stop Smoking group<br>6. Attended one or more Stop Smoking one-to-one counselling\advice\support session\s<br>7. Phoned a Smoking Helpline<br>8. Visited www.nhs.uk\smokefree website<br>9. Visited a website other than Smokefree<br>10. Used an application ('app') on a handheld computer (smartphone, tablet, PDA)<br>11. Hypnotherapy<br>12. Acupuncture<br>13. Electronic cigarette<br>14. Heat-not-burn cigarette (e.g. iQOS, heatsticks)<br>15. Juul<br>16. Allen Carr Easyway session<br>17. Allen Carr Easyway book<br>18. The SmokeFree Formula book<br>19. Other book or booklet<br>20. Tobacco-free nicotine pouch/pod or 'white pouches'<br>21. Cytisine (e.g. Tabex, Tactizen or Desmoxan)<br>22. Other (please specify) |

| | | |
|---|---|---|
| | 16. Used an alternative therapy (for example, hypnotherapy, acupuncture, homeopathy, Chinese medicine)<br>17. Signed up for a Pharmacy stop smoking programme (for example, Boots 'Change One Thing', Lloyds, Tesco)<br>18. Used a non-NHS programme, such as Allen Carr<br>19. Talked about stopping with family and friends<br>20. Joined a social media group (for example, on Facebook)<br>21. Used a local Stop Smoking Service (face to face or online)<br>22. Other (please specify)<br>23. Used willpower alone<br>24. None of these<br>25. Don't know | |
| Coding: digital support | 8-13 = yes<br>else = no | 8-10 = yes<br>else = no |
| Coding: stop smoking services | 21 = yes<br>else = no | 5-6 = yes<br>else = no |

**Success of quit attempts**

| Base: adults who smoked in the past year and made at least one past-year quit attempt | | |
|---|---|---|
| Survey | **Ipsos pre/post campaign** | **Smoking Toolkit Study** |
| Variable name | F1 | Q632b9 |
| Item wording and response options | Have you made an attempt to quit smoking in the past 12 months?<br>By quit attempt, we mean stopping smoking cigarettes or other tobacco products (e.g. pipe, cigar, shisha, etc.), though you may still be using a vape (electronic cigarette or vaping device).<br><br>1. Yes - and I am still not smoking cigarettes<br>2. Yes - but I have started smoking cigarettes again<br>3. No - I have not made a quit attempt<br>4. Don't know | How long did your most recent serious quit attempt last before<br>you went back to smoking?<br><br>1. Still not smoking<br>2. Less than a day<br>3. Less than a week<br>4. More than 1 week and up to a month<br>5. More than 1 month and up to 2 months<br>6. More than 2 months and up to 3 months<br>7. More than 3 months and up to 6 months<br>8. More than 6 months and up to a year |
| Coding: quit success | 1 = yes<br>2 = no<br>3-4 = NA | 1 = yes<br>2-8 = no |
