## Supplementary file 2 for "Are population-wide mass media campaigns still relevant in an era of low smoking prevalence? Insights from the *Power to Quit* campaign in England"

**Table S1.** Weighted sample characteristics, pre- and post-campaign surveys commissioned by the Department for Health and Social Care

| | Adults who smoked in the past year | | | Adults who attempted to quit smoking in the past year | | |
|---|---|---|---|---|---|---|
| | **Pre** | **Post (all)** | **Post (recognised campaign materials)** | **Pre** | **Post (all)** | **Post (recognised campaign materials)** |
| Unweighted *n* | 803 | 1,713 | 799 | 383 | 901 | 518 |
| Age (years), % | | | | | | |
| 18-24 | 9.3 | 14.7 | 18.0 | 11.1 | 16.3 | 19.5 |
| 25-34 | 37.0 | 33.9 | 41.9 | 40.2 | 39.1 | 45.1 |
| 35-44 | 16.7 | 17.0 | 16.5 | 18.3 | 16.4 | 15.2 |
| 45-54 | 18.5 | 16.8 | 11.6 | 15.9 | 14.9 | 10.3 |
| 55-64 | 12.7 | 11.6 | 8.3 | 10.3 | 9.1 | 7.6 |
| ≥65 | 5.7 | 6.2 | 3.7 | 4.2 | 4.4 | 2.4 |
| Gender, % | | | | | | |
| Man | 58.9 | 58.2 | 60.8 | 57.0 | 58.0 | 60.2 |
| Woman | 40.5 | 40.9 | 38.2 | 42.1 | 41.1 | 38.3 |
| Other | 0.6 | 0.9 | 1.0 | 1.0 | 0.9 | 1.5 |
| Occupational social grade, % | | | | | | |
| ABC1 (more advantaged) | 54.1 | 55.9 | 58.6 | 57.6 | 56.6 | 59.6 |
| C2DE (less advantaged) | 45.9 | 44.1 | 41.4 | 42.4 | 43.4 | 40.4 |

Pre, pre-campaign survey (4–11 February 2026).
Post, post-campaign survey (9–23 April 2026).

**Table S2.** Weighted sample characteristics, Smoking Toolkit Study

| | **Adults who smoked in the past year** | **Adults who attempted to quit smoking in the past year** |
|---|---|---|
| Unweighted *n* | 11,442 | 4,091 |
| Age (years), % | | |
| 18-24 | 16.9 | 21.9 |
| 25-34 | 23.8 | 27.4 |
| 35-44 | 18.1 | 17.5 |
| 45-54 | 15.2 | 12.7 |
| 55-64 | 14.0 | 12.0 |
| ≥65 | 12.0 | 8.5 |
| Gender, % | | |
| Man | 53.5 | 52.5 |
| Woman | 45.0 | 46.1 |
| Other | 1.5 | 1.4 |
| Occupational social grade, % | | |
| ABC1 (more advantaged) | 42.7 | 42.1 |
| C2DE (less advantaged) | 57.3 | 57.9 |

Data collected January 2023 – June 2026.

**Table S3.** Prevalence of quit attempts over time, overall and by occupational social grade

| | Quit attempt, % [95% CI] | | | | | | |
|---|---|---|---|---|---|---|---|
| | **Jan-June 2023** | **July-Dec 2023** | **Jan-June 2024** | **July-Dec 2024** | **Jan-June 2025** | **July-Dec 2025** | **Jan-June 2026** |
| **Adults who smoked in the past year** | | | | | | | |
| Overall | 38.1 [35.6–40.7] | 34.0 [31.6–36.5] | 37.6 [35.0–40.2] | 37.6 [34.9–40.3] | 36.9 [34.4–39.3] | 33.7 [31.2–36.1] | 36.6 [33.9–39.2] |
| ABC1 (more advantaged) | 36.6 [33.4–39.9] | 33.4 [30.2–36.6] | 37.5 [34.2–40.9] | 37.6 [34.3–40.8] | 35.1 [32.0–38.1] | 34.5 [31.5–37.6] | 36.0 [32.7–39.4] |
| C2DE (less advantaged) | 39.3 [35.5–43.2] | 34.5 [30.9–38.1] | 37.7 [34.0–41.4] | 37.6 [33.6–41.7] | 38.2 [34.6–41.8] | 33.0 [29.3–36.7] | 36.9 [33.1–40.8] |

CI, confidence interval. SSS, Stop Smoking Services.
Data shown are unmodelled weighted prevalence estimates from the Smoking Toolkit Study, aggregated in six-month periods.

**Table S4.** Prevalence of quit attempts involving digital support over time, overall and by occupational social grade

| | Quit attempt involving digital support, % [95% CI] | | | | | | |
|---|---|---|---|---|---|---|---|
| | Jan-June 2023 | July-Dec 2023 | Jan-June 2024 | July-Dec 2024 | Jan-June 2025 | July-Dec 2025 | Jan-June 2026 |
| **Adults who smoked in the past year** | | | | | | | |
| Overall | 1.7 [1.0–2.5] | 2.2 [1.4–3.0] | 3.4 [2.4–4.4] | 3.1 [2.1–4.1] | 2.9 [2.0–3.8] | 3.2 [2.3–4.2] | 5.1 [3.9–6.4] |
| ABC1 (more advantaged) | 1.4 [0.6–2.1] | 1.5 [0.7–2.3] | 3.6 [2.2–4.9] | 3.0 [1.9–4.1] | 3.1 [2.0–4.2] | 3.0 [1.9–4.2] | 5.1 [3.5–6.7] |
| C2DE (less advantaged) | 2.1 [0.9–3.2] | 2.7 [1.5–3.9] | 3.3 [2.0–4.7] | 3.2 [1.7–4.8] | 2.7 [1.5–4.0] | 3.4 [1.9–4.9] | 5.2 [3.4–7.0] |
| **Adults who attempted to quit smoking in the past year** | | | | | | | |
| Overall | 4.6 [2.7–6.4] | 6.5 [4.3–8.7] | 9.1 [6.6–11.6] | 8.3 [5.7–10.9] | 7.9 [5.6–10.1] | 9.6 [6.9–12.4] | 14.1 [10.8–17.3] |
| ABC1 (more advantaged) | 3.7 [1.7–5.8] | 4.5 [2.2–6.8] | 9.5 [6.0–13.0] | 7.9 [5.0–10.8] | 8.9 [5.8–12.0] | 8.8 [5.6–12.0] | 14.1 [10.0–18.3] |
| C2DE (less advantaged) | 5.2 [2.4–8.1] | 7.9 [4.5–11.3] | 8.8 [5.3–12.3] | 8.6 [4.7–12.5] | 7.1 [3.9–10.3] | 10.3 [6.0–14.5] | 14.0 [9.4–18.6] |

CI, confidence interval. SSS, Stop Smoking Services.
Data shown are unmodelled weighted prevalence estimates from the Smoking Toolkit Study, aggregated in six-month periods.

**Table S5.** Prevalence of quit attempts involving local Stop Smoking Services over time, overall and by occupational social grade

| | **Quit attempt involving local Stop Smoking Services, % [95% CI]** | | | | | | |
|---|---|---|---|---|---|---|---|
| | **Jan-June 2023** | **July-Dec 2023** | **Jan-June 2024** | **July-Dec 2024** | **Jan-June 2025** | **July-Dec 2025** | **Jan-June 2026** |
| **Adults who smoked in the past year** | | | | | | | |
| Overall | 0.7 [0.3–1.1] | 0.6 [0.2–1.1] | 1.2 [0.7–1.8] | 1.6 [1.0–2.3] | 1.4 [0.9–1.9] | 2.3 [1.5–3.1] | 3.8 [2.8–4.9] |
| ABC1 (more advantaged) | 0.6 [0.1–1.1] | 0.4 [0.0–0.9] | 0.8 [0.2–1.4] | 1.5 [0.7–2.3] | 1.6 [0.9–2.4] | 1.1 [0.5–1.8] | 3.4 [2.1–4.7] |
| C2DE (less advantaged) | 0.7 [0.1–1.3] | 0.8 [0.1–1.4] | 1.5 [0.7–2.3] | 1.7 [0.7–2.7] | 1.2 [0.4–1.9] | 3.2 [1.9–4.5] | 4.2 [2.6–5.7] |
| **Adults who attempted to quit smoking in the past year** | | | | | | | |
| Overall | 1.8 [0.7–2.8] | 1.9 [0.6–3.1] | 3.3 [1.8–4.7] | 4.3 [2.6–6.1] | 3.8 [2.3–5.2] | 6.8 [4.5–9.1] | 10.5 [7.7–13.3] |
| ABC1 (more advantaged) | 1.7 [0.3–3.0] | 1.3 [0.0–2.6] | 2.2 [0.5–3.8] | 4.0 [1.9–6.0] | 4.7 [2.6–6.8] | 3.2 [1.3–5.1] | 9.3 [5.9–12.8] |
| C2DE (less advantaged) | 1.8 [0.4–3.3] | 2.3 [0.4–4.2] | 4.0 [1.9–6.2] | 4.6 [1.9–7.2] | 3.1 [1.2–5.0] | 9.8 [5.9–13.6] | 11.3 [7.2–15.3] |

CI, confidence interval. SSS, Stop Smoking Services.
Data shown are unmodelled weighted prevalence estimates from the Smoking Toolkit Study, aggregated in six-month periods.

**Table S6.** Prevalence of quit success involving digital support over time, overall and by occupational social grade

| | Quit success, % [95% CI] | | | | | | |
|---|---|---|---|---|---|---|---|
| | **Jan-June 2023** | **July-Dec 2023** | **Jan-June 2024** | **July-Dec 2024** | **Jan-June 2025** | **July-Dec 2025** | **Jan-June 2026** |
| **Adults who smoked in the past year** | | | | | | | |
| Overall | 9.7 [8.1–11.3] | 8.0 [6.7–9.4] | 10.0 [8.5–11.6] | 10.4 [8.7–12.1] | 10.9 [9.3–12.5] | 9.4 [7.9–10.9] | 12.8 [10.9–14.7] |
| ABC1 (more advantaged) | 8.9 [7.0–10.7] | 9.5 [7.5–11.5] | 11.9 [9.6–14.2] | 12.9 [10.6–15.2] | 11.0 [9.1–13.0] | 11.2 [9.1–13.2] | 12.4 [10.1–14.7] |
| C2DE (less advantaged) | 10.3 [7.9–12.8] | 6.9 [5.1–8.8] | 8.8 [6.6–10.9] | 8.5 [6.2–10.9] | 10.8 [8.4–13.1] | 8.0 [5.9–10.1] | 13.1 [10.3–15.9] |
| **Adults who attempted to quit smoking in the past year** | | | | | | | |
| Overall | 25.4 [21.6–29.2] | 23.6 [19.9–27.3] | 26.7 [22.9–30.5] | 27.7 [23.6–31.7] | 29.6 [25.7–33.4] | 27.9 [23.9–31.9] | 35.0 [30.5–39.5] |
| ABC1 (more advantaged) | 24.2 [19.6–28.9] | 28.5 [23.2–33.8] | 31.7 [26.4–37.1] | 34.3 [29.1–39.6] | 31.5 [26.5–36.5] | 32.3 [27.1–37.5] | 34.4 [28.8–40.0] |
| C2DE (less advantaged) | 26.3 [20.6–31.9] | 20.1 [15.0–25.1] | 23.3 [18.0–28.5] | 22.7 [16.9–28.5] | 28.2 [22.7–33.6] | 24.4 [18.6–30.2] | 35.4 [28.9–41.8] |

CI, confidence interval. SSS, Stop Smoking Services.
Data shown are weighted prevalence estimates from the Smoking Toolkit Study, aggregated in six-month periods.
